# Clinical Risk Factors and a Prediction Model for Placenta Accreta Spectrum Among Women Without Prior Cesarean Delivery: A Single-Center Cohort Study

**DOI:** 10.64898/2026.05.30.26354499

**Authors:** Xiaotang Zhai, Huanyu You, Jiyu Wei, Nan Wang, Lin Zeng, Yangyu Zhao, Yan Wang

**Affiliations:** Department of Obstetrics and Gynecology, Peking University Third Hospital, Beijing 100191, China; National Centre for Healthcare Quality Management in Obstetrics, Beijing, China; Research Center of Clinical Epidemiology, Peking University Third Hospital, Beijing, China

**Keywords:** placenta accreta spectrum, prediction model, cohort study, cesarean section, operative hysteroscopy, risk factors, placenta previa

## Abstract

**Background:** Placenta accreta spectrum (PAS) is an important cause of severe maternal morbidity. Although prior cesarean delivery is a well-established risk factor, PAS also occurs in women without prior cesarean section (CS), in whom risk may be underestimated. This study evaluated routinely available clinical factors associated with PAS in this population and developed a clinical-history-based prediction model.

**Methods:** We conducted a retrospective cohort study of women without prior CS who delivered at Peking University Third Hospital, China, from January 1, 2022, to December 31, 2023. PAS was diagnosed according to the 2019 International Federation of Gynecology and Obstetrics clinical and/or histopathological criteria. Multivariable logistic regression was used to identify independent risk factors. Model performance was assessed using receiver operating characteristic curves, calibration, decision curve analysis, and stratified 5-fold cross-validation. Analyses were repeated after stratification by placenta previa status.

**Results:** Among 11,148 women without prior CS, 236 had PAS. Independent risk factors in the overall cohort were placenta previa, operative hysteroscopy, uterine curettage, in vitro fertilization, and multifetal pregnancy. The overall clinical prediction model showed an area under the curve of 0.838 (95% confidence interval, 0.81-0.87), with stable performance in internal validation. In stratified analyses, model discrimination was lower among women without placenta previa (area under the curve, 0.734) and those with placenta previa (area under the curve, 0.647).

**Conclusions:** In this single-center cohort, routinely available clinical history was associated with PAS risk among women without prior CS. The proposed model may help identify patients who warrant targeted PAS imaging or specialist assessment, but external validation and integration with imaging features are needed before broad clinical implementation.

## Introduction

Placenta accreta spectrum (PAS) disorders, a spectrum of conditions characterized by abnormal invasion of placental trophoblasts into the myometrium, have seen a rising incidence in recent years[1, 2]. PAS is a leading cause of severe postpartum hemorrhage, peripartum hysterectomy, and maternal mortality[3]. Previous studies indicated that the highest-risk population for PAS comprises patients with a prior cesarean section (CS) and coexisting placenta previa[1]. However, a nationwide study in United States revealed that approximately one-third of PAS cases occur in patients without prior CS[4]. Currently, the identification strategy for high-risk populations of PAS in patients without prior CS remains unclear, potentially leading to underdiagnosis and missed opportunities for delivery planning and outcome optimization. Patients without prior CS warrant particular attention; however, most current studies focused on the general population.

Previous studies have primarily examined single exposure factors in relation to PAS risk, such as hysteroscopic adhesiolysis, in vitro fertilization (IVF), uterine malformation, adenomyosis, twin pregnancy, and myomectomy [5–11]. In a large study of primiparous women, intrauterine procedures were independently associated with PAS, and the risk increased with the cumulative number of procedures [12]. In our previous case-control study of women without prior CS, operative hysteroscopy and uterine curettage were associated with PAS after adjustment for placenta previa; that study also highlighted the importance of classifying and quantifying intrauterine surgical history [13]. However, cohort-based estimates and prediction models specifically focused on women without prior CS remain limited. The present study therefore aimed to evaluate clinical risk factors for PAS in a defined delivery cohort of women without prior CS and to develop a transparent, clinical-history-based prediction model for risk stratification.

## Materials and Methods

### Study design and population

We retrospectively enrolled all women without prior cesarean delivery who delivered at Peking University Third Hospital between January 1, 2022, and December 31, 2023 (n = 11,148). The study was approved by the Ethics Committee of Peking University Third Hospital (IRB00006761-M2022827), and the requirement for informed consent was waived because of the retrospective design and use of routinely collected clinical data. Authors had no access to information that could identify individual participants during or after data collection.

### Maternal characteristics

Maternal characteristics were obtained from our center’s electronic medical record system or telephone follow-up. Complete data were available for the variables included in the final analyses. Maternal characteristics included maternal age at delivery; pre-pregnancy body mass index (BMI); parity; prior and current pregnancy history, including IVF, multifetal pregnancy, gestational diabetes mellitus, hypertensive disorders of pregnancy, and placenta previa; medical history, including pre-gestational diabetes mellitus and autoimmune disease; intrauterine surgery; laparoscopic or open uterine surgery; and gynecological history, including congenital uterine malformation, uterine myomas, or adenomyosis. Autoimmune diseases included systemic lupus erythematosus, Sjögren syndrome, rheumatoid arthritis, ankylosing spondylitis, Behçet disease, confirmed antiphospholipid syndrome, and polymyositis. Placenta previa was defined as placenta overlying the internal os to any degree. Intrauterine surgeries were categorized into operative hysteroscopy (including adhesiolysis, uterine septum resection, and uterine myomectomy), uterine curettage, and endometrial biopsy (including endometrial biopsy, simple hysteroscopy, and endometrial polypectomy).

### Outcome measures

The diagnosis of PAS served as the outcome variable. PAS diagnosis followed the 2019 International Federation of Gynecology and Obstetrics (FIGO) clinical and/or histopathological diagnostic criteria [14]. Diagnosis was confirmed by intraoperative findings and/or pathological examination in cesarean delivery patients, and by clinical diagnostic criteria in vaginal delivery patients.

### Statistical analysis

Statistical analyses were performed using SPSS 27.0 (SPSS Inc., Armonk, NY, USA), with categorical variables analyzed by chi-square or Fisher’s exact tests and continuous variables compared using Student’s t-tests. Binary logistic regression models were employed to adjust for multiple variables.

R version 4.3.1 (R Foundation for Statistical Computing, Vienna, Austria) was used for model construction and validation. The “rms” package was used for nomogram construction and calibration; “rmda” for decision curve analysis; “ResourceSelection” for the Hosmer-Lemeshow test; and “pROC” for receiver operating characteristic (ROC) curve analysis. All tests were two-tailed, and P < 0.05 was considered statistically significant. The reporting of this observational prediction model study was guided by the STROBE and TRIPOD recommendations where applicable.

To assess internal model stability, internal validation was conducted using stratified 5-fold cross-validation, which preserved the overall proportion of positive and negative samples in each fold. Model performance was evaluated using the mean area under the curve (AUC), 95% confidence interval (CI), and standard deviation (SD).

## Results

This study included 11,148 women without prior CS who gave birth at Peking University Third Hospital between January 1, 2022, and December 31, 2023. Among these, 236 cases meeting the 2019 FIGO clinical criteria for PAS were enrolled in the PAS group, while the remaining 10,912 women comprised the non-PAS group (Figure 1). Stratified analyses were conducted according to placenta previa status. The cohort comprised 345 women with placenta previa and 10,803 women without placenta previa (Figure 1). Based on clinical and/or pathological diagnostic criteria, the prevalence of PAS was 2.1% (236/11,148) among women without prior CS, 1.2% (132/10,803) among women with neither prior CS nor placenta previa, and 30.1% (104/345) among women without prior CS but with placenta previa.

**Figure 1.**
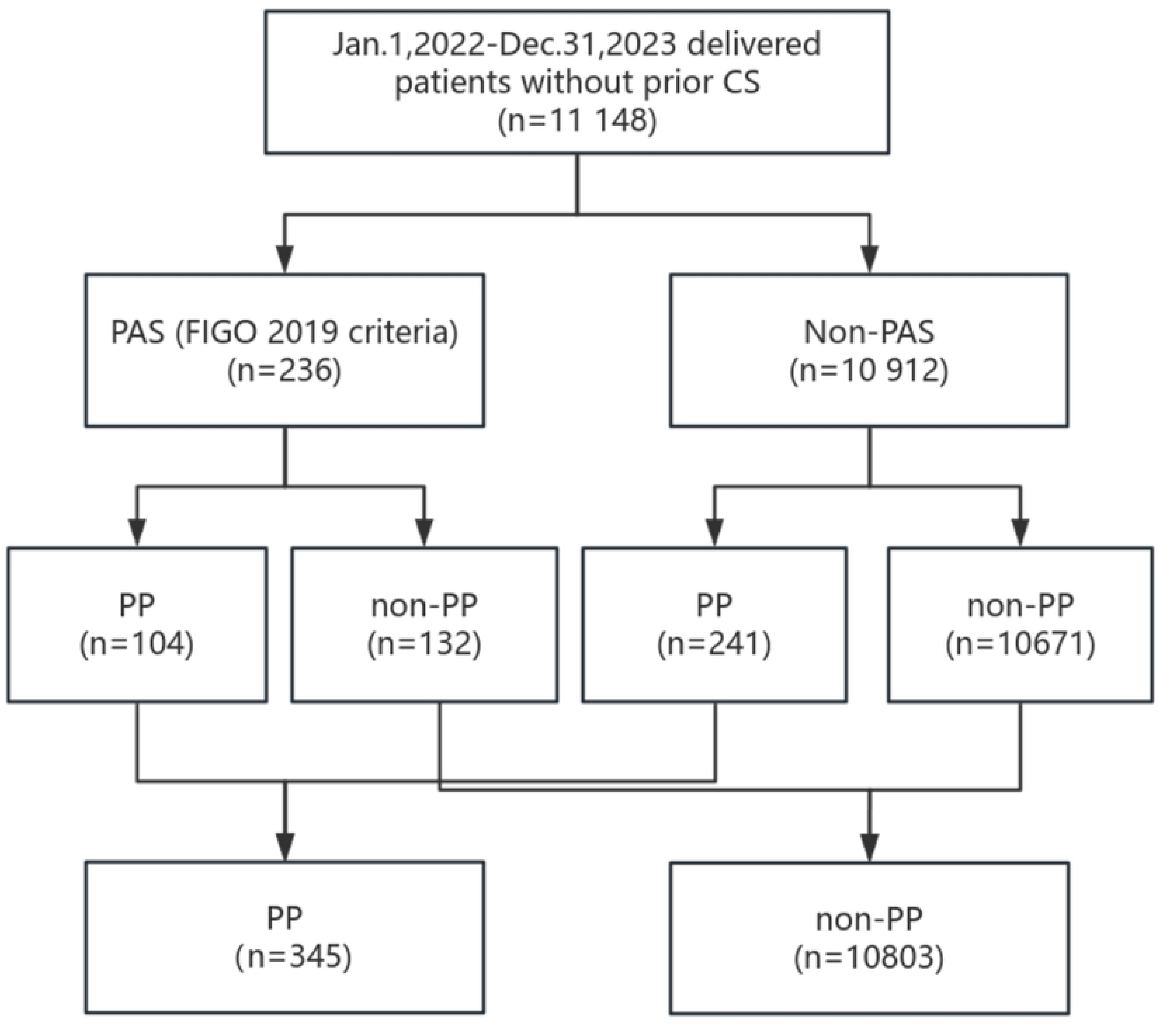
Flow diagram of included patients in the study and control groups. CS, cesarean section, PAS, placenta accreta spectrum. PP, placenta previa.

### Patients without prior CS

Analysis of risk factors for PAS in women without prior CS revealed significant differences between groups. Compared with the non-PAS group, the PAS group had higher proportions of advanced maternal age, IVF, adenomyosis, uterine malformation, multifetal pregnancy, placenta previa, and intrauterine surgery (Table 1). Among the 236 PAS patients, 177 (75.3%) were diagnosed with placenta accreta, 51 (21.6%) with placenta increta, and 7 (3.0%) with placenta percreta. Vaginal delivery occurred in 37 cases (15.7%), while cesarean delivery was performed in 199 cases (84.3%). Therapeutic hysterectomy was performed in 2 patients with placenta percreta, with histopathological examination confirming PAS in both cases. Placental pathology also supported the diagnosis of PAS in 12 patients.

**Table 1.**
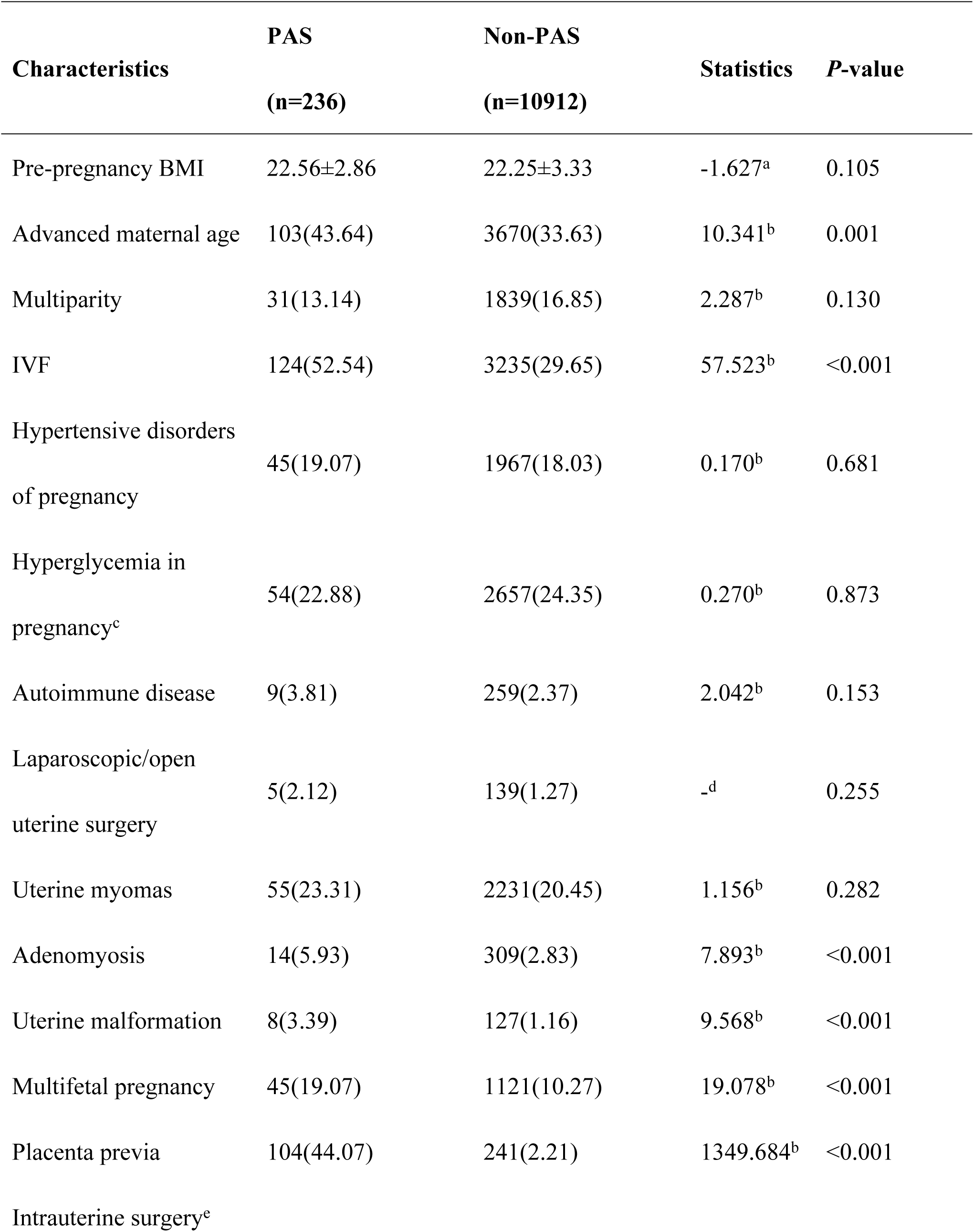
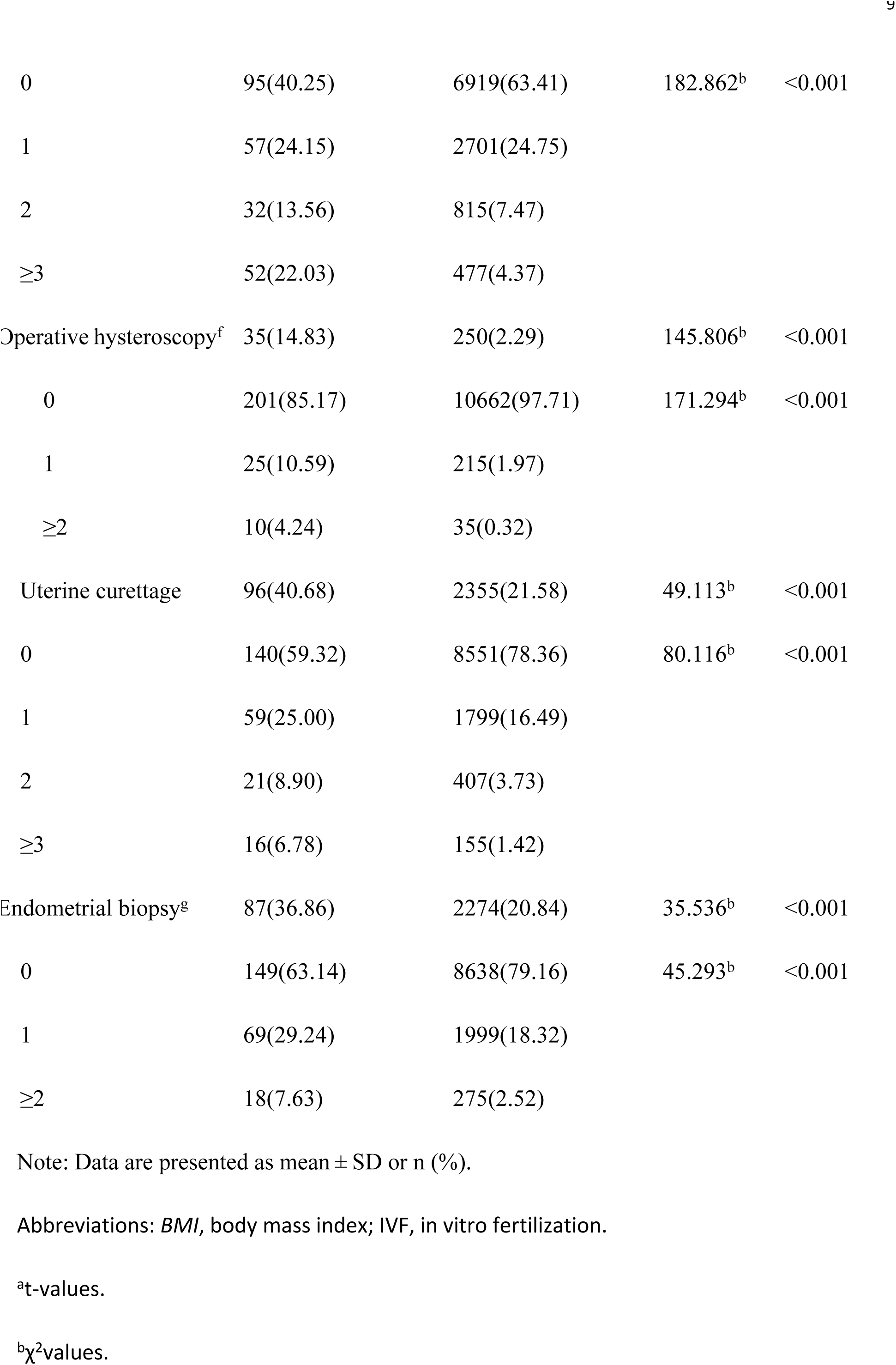
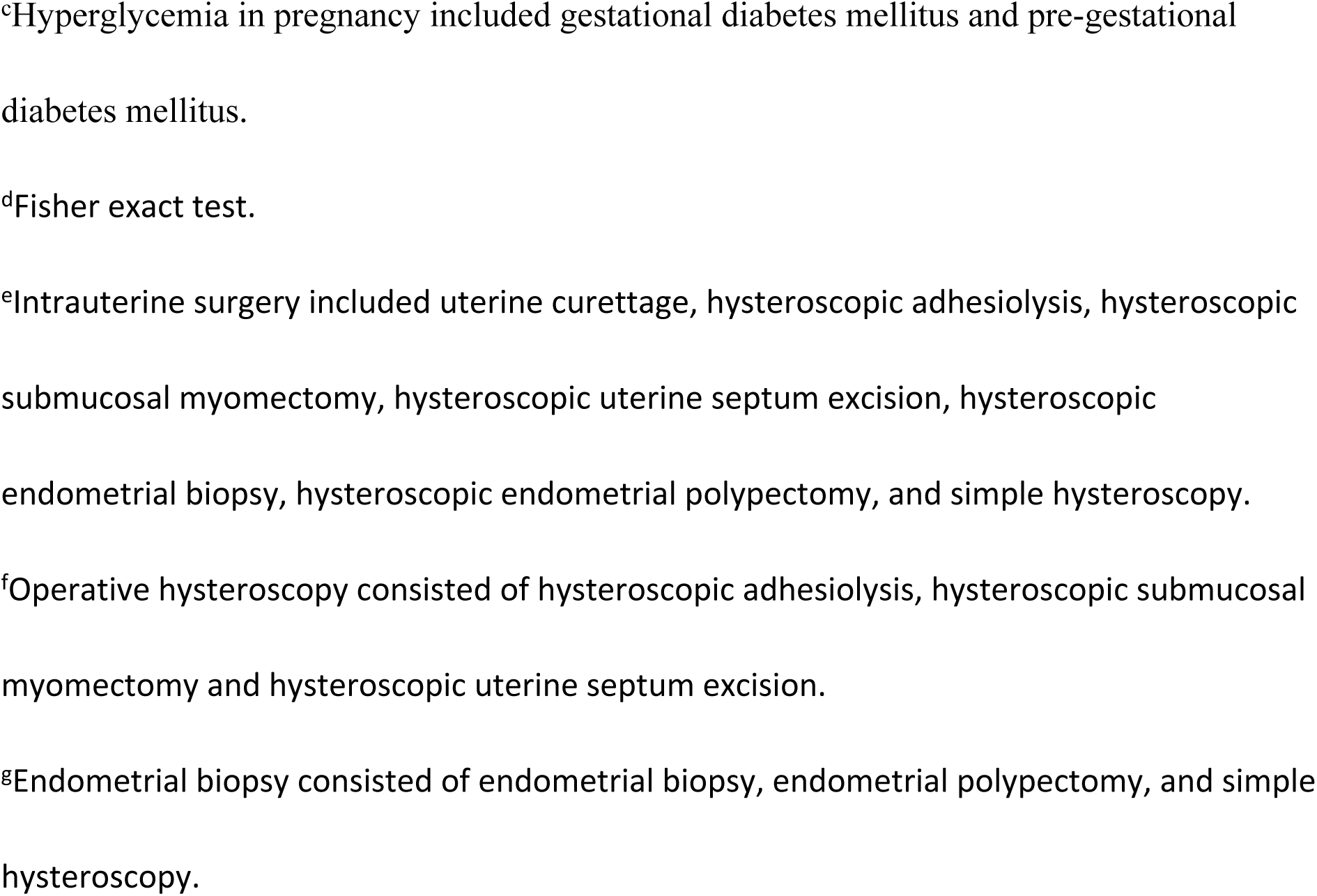
Baseline characteristics in placenta accreta spectrum (PAS) patients without prior cesarean section (CS) and non-PAS controls.

Since different types and frequencies of intrauterine surgeries may cause varying degrees of endometrial damage, this study collected detailed medical histories and conducted statistical analyses, as shown in Table 1. Patients in the PAS group had a higher proportion and more times of uterine curettage, endometrial biopsy, and operative hysteroscopy.

Binary logistic regression was used to adjust for confounding factors. Variables with P < 0.1 in univariate analysis and multiparity, a previously reported risk factor, were included in the multivariable model [13]. Operative hysteroscopy (once: aOR 3.74, 95% CI 2.15-6.49, P < 0.001; two or more times: aOR 6.47, 95% CI 2.46-17.04, P < 0.001), uterine curettage (once: aOR 1.74, 95% CI 1.23-2.47, P = 0.002; twice: aOR 2.30, 95% CI 1.33-3.98, P = 0.003; three or more times: aOR 5.04, 95% CI 2.64-9.61, P < 0.001), IVF (aOR 1.75, 95% CI 1.26-2.43, P = 0.001), multifetal pregnancy (aOR 1.56, 95% CI 1.06-2.30, P = 0.026), and placenta previa (aOR 34.15, 95% CI 25.09-46.49, P < 0.001) were independently associated with PAS (Table 2).

**Table 2.**
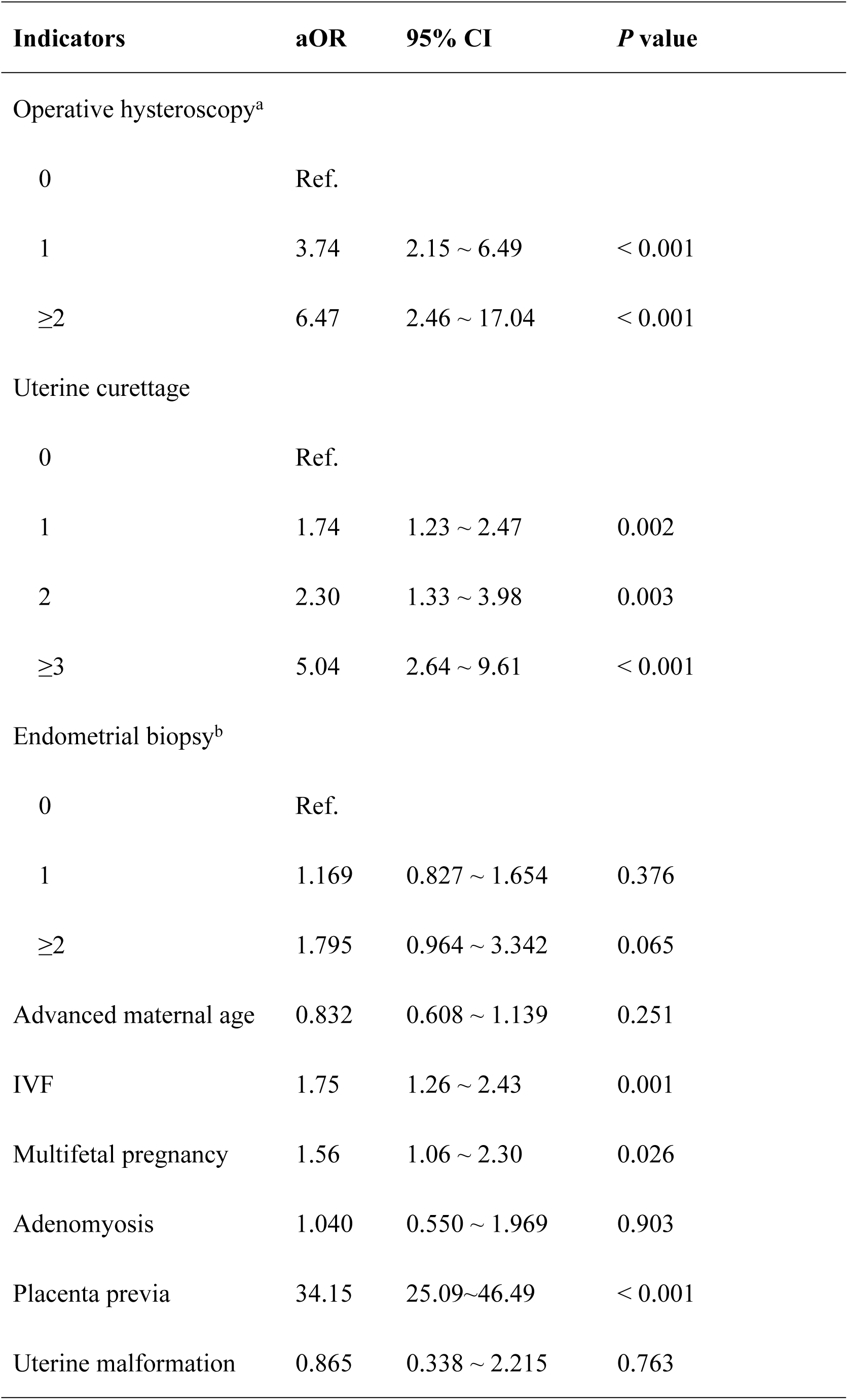
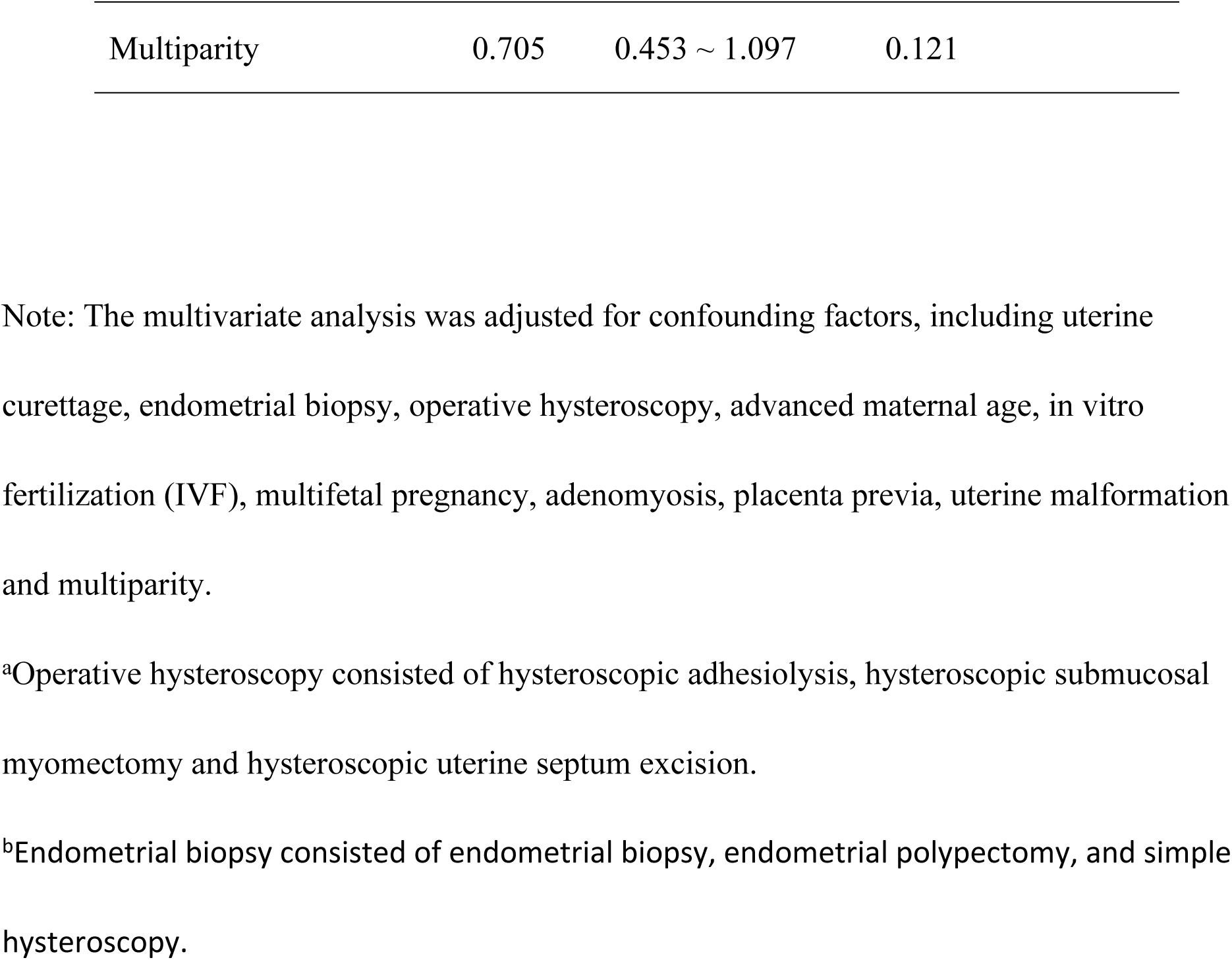
Multivariate analysis of risk factors for placenta accreta spectrum (PAS) patients without prior cesarean section (CS).

The regression model passed the Hosmer-Lemeshow test (P = 0.102). Based on the final multivariable logistic regression model, a nomogram was constructed to estimate PAS risk among women without prior CS (Figure 2). The ROC curve of the nomogram (Figure 3a) showed an AUC of 0.838 (95% CI 0.81-0.87), with a cutoff value of 0.019. The calibration curve (Figure 3b) indicated better agreement between predicted and observed PAS risk in lower-risk ranges, while overestimation may occur in higher-risk ranges. Decision curve analysis (Figure 3c) showed positive net benefit across clinically relevant threshold ranges. At the ROC-derived cutoff value, sensitivity was 0.712, specificity was 0.832, positive predictive value (PPV) was 0.084, and negative predictive value (NPV) was 0.993. The nomogram was generated from the coefficients of the final logistic regression model.

**Figure 2.**
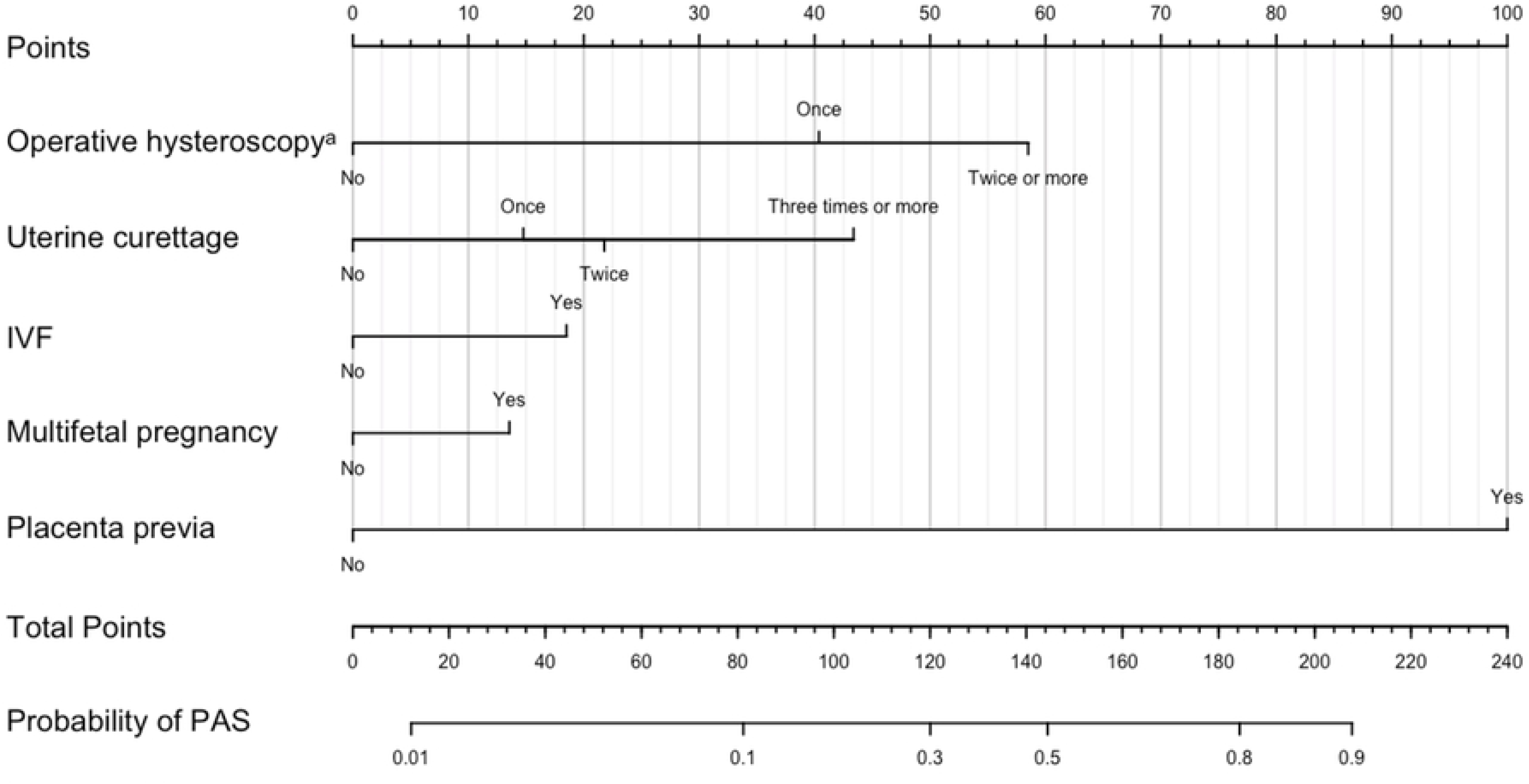
Nomogram to estimate the risk of placenta accreta spectrum (PAS) in patients without prior cesarean section (CS). IVF, in vitro fertilization. ^a^Operative hysteroscopy consisted of hysteroscopic adhesiolysis, hysteroscopic submucosal myomectomy and hysteroscopic uterine septum excision.

**Figure 3.**
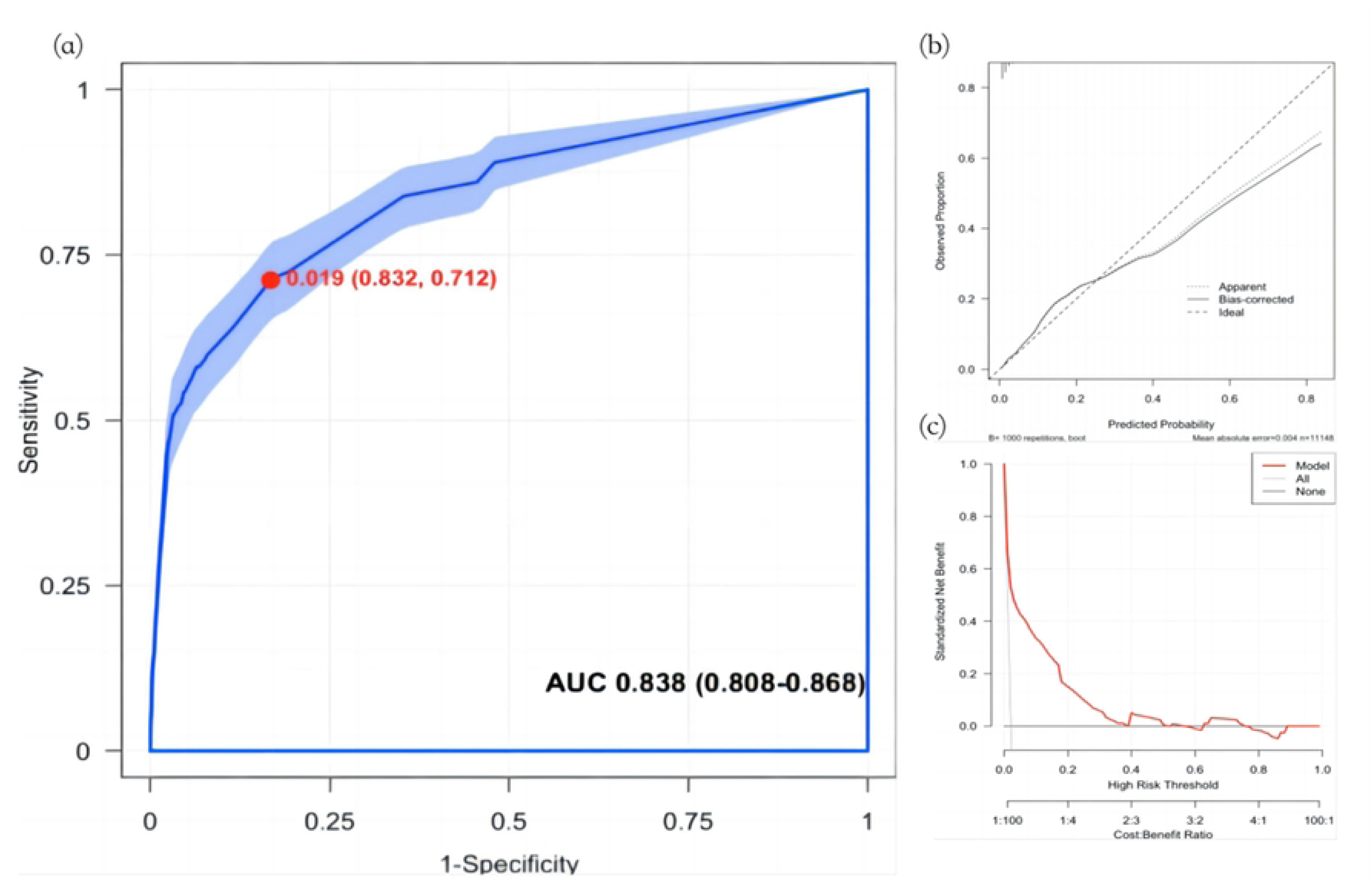
(a) Receiver operating characteristic curve (ROC curve) of the clinical prediction model. The cut-off value (95%CI) was marked on the curve and area under the curve (AUC) (95%CI) was marked below. (b) Calibration curves. (c) Decision curve. Analysis evaluating model clinical utility, where curves above the ‘None’ line and the ‘All’ line indicate positive net benefit at corresponding risk thresholds.

Internal validation using stratified 5-fold cross-validation showed a mean AUC of 0.837 (95% CI 0.78-0.87), with an SD of 0.040, suggesting stable internal performance (Supplemental Figure 1a).

### Patients with neither prior CS nor placenta previa

Placenta previa is an established independent risk factor for PAS [1] and demonstrated the strongest association in this study (aOR 34.15, 95% CI 25.09-46.49). Stratified analysis by placenta previa status was therefore conducted (Figure 1). Supplemental Table 1 shows the baseline characteristics of women with neither prior CS nor placenta previa. A binary logistic regression model constructed using factors with P < 0.1 in the univariate analysis (Supplemental Table 2) showed that uterine curettage (once: aOR 1.68, 95% CI 1.10-2.58, P = 0.017; twice or more: aOR 2.66, 95% CI 1.54-4.59, P < 0.001), operative hysteroscopy (aOR 3.92, 95% CI 2.23-6.88, P < 0.001), IVF (aOR 2.30, 95% CI 1.52-3.47, P < 0.001), and multifetal pregnancy (aOR 2.13, 95% CI 1.40-3.24, P < 0.001) were independently associated with PAS in women with neither prior CS nor placenta previa. The regression model passed the Hosmer-Lemeshow test (P = 0.636).

Based on the results of the binary logistic regression analysis, this study combined independent risk factors to construct a nomogram (Figure 4a). The ROC curve of the nomogram showed an AUC of 0.734 (95% CI 0.69–0.78). Using the cutoff value from the ROC curve as the clinical decision-making threshold for this model yielded a sensitivity of 0.765, specificity of 0.636, PPV of 0.025, and NPV of 0.995.

**Figure 4.**
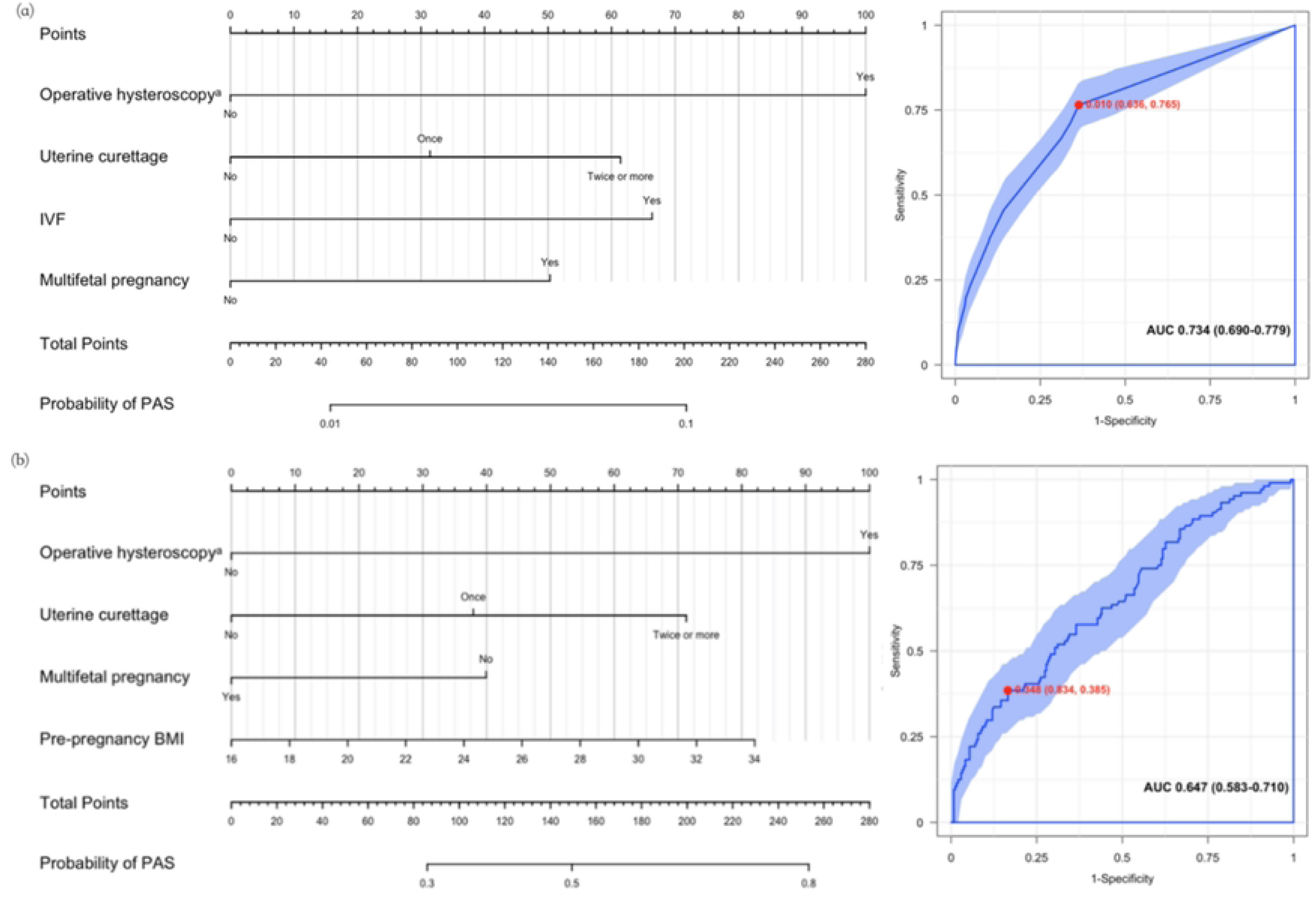
Nomogram and receiver operating characteristic curve (ROC curve) to estimate the risk of placenta accreta spectrum (PAS) in patients (a) with neither prior cesarean section (CS) nor placenta previa and (b) without prior cesarean section (CS) but with placenta previa. The cutoff value (95% CI) was marked on the ROC curve, and area under the curve (AUC) (95% CI) was marked below. ^a^Operative hysteroscopy consisted of hysteroscopic adhesiolysis, hysteroscopic submucosal myomectomy and hysteroscopic uterine septum excision.

Internal validation showed a mean AUC of 0.695 (95% CI 0.66-0.74), with an SD of 0.033, indicating moderate internal performance in this subgroup (Supplemental Figure 1b).

### Patients without prior CS but with placenta previa

#Supplemental Table 3 shows the baseline characteristics of women without prior CS but with placenta previa. The binary logistic regression model included variables with P < 0.2 in univariate analysis, including uterine curettage, operative hysteroscopy, pre-pregnancy BMI, and multifetal pregnancy (Supplemental Table 4). Laparoscopic/open uterine surgery was not included due to the insufficient number of cases. Uterine curettage (aOR 2.59, 95% CI 1.19-5.62, P = 0.016) and operative hysteroscopy (aOR 3.80, 95% CI 1.43-10.07, P = 0.007) were independently associated with PAS in this subgroup. This model passed the Hosmer-Lemeshow test (P = 0.857). A nomogram was constructed using operative hysteroscopy, uterine curettage, pre-pregnancy BMI, and multifetal pregnancy (Figure 4b). The ROC curve showed an AUC of 0.647 (95% CI 0.58-0.71). At the ROC-derived cutoff value, sensitivity was 0.385, specificity was 0.834, PPV was 0.500, and NPV was 0.758. Internal validation showed a mean AUC of 0.629 (95% CI 0.60-0.672), with an SD of 0.032 (Supplemental Figure 1c). Diagnostic performance metrics for the three predictive models are summarized in Supplemental Table 5.

## Discussion

In this single-center cohort of 11,148 women without prior CS, placenta previa, operative hysteroscopy, uterine curettage, IVF, and multifetal pregnancy were independently associated with PAS. Detailed classification of intrauterine surgical history, including type and frequency of procedures, provided clinically useful risk information. We also developed and internally validated a clinical-history-based prediction model. The model is intended to support risk stratification and selection of patients for targeted PAS imaging or specialist assessment, rather than to replace diagnostic ultrasound or multidisciplinary clinical judgment.

Placenta previa (aOR 34.15) remained the strongest independent risk factor for PAS in this cohort, consistent with previous studies [1, 4, 15, 16]. In a prior large cohort study of primiparous women, placenta previa demonstrated a relative risk of 6.2 (95% CI 4.4-8.8) [12]. Our findings support the continued importance of placenta previa in PAS risk assessment among women without prior CS. However, precise risk stratification in this population requires attention to additional clinical history and imaging findings.

Operative hysteroscopy (once: aOR 3.74; two or more times: aOR 6.47) and uterine curettage (once: aOR 1.74; twice: aOR 2.30; three or more times: aOR 5.04) were independently associated with PAS, with higher estimates observed after repeated procedures. Endometrial biopsy was not independently associated with PAS after adjustment, which is consistent with our earlier case-control study of women without prior CS [13]. These results are also broadly consistent with previous studies [1, 12], although earlier reports often lacked detailed classification of procedure type and frequency. Intrauterine procedures may contribute to PAS through endometrial and decidual injury [17], but our data suggested that only substantial endometrial injury triggers sufficient dysfunction to initiate PAS.. Clinically, careful documentation of intrauterine surgical history may improve risk assessment.

IVF (aOR 1.75) and multifetal pregnancy (aOR 1.56) were also independently associated with PAS among women without prior CS, consistent with previous studies [8, 18]. IVF may be associated with endometrial factors relevant to abnormal placentation, and studies have linked ovarian stimulation protocols and endometrial thickness to PAS risk [5, 19]. The association between multifetal pregnancy and PAS remains incompletely understood, but a larger placental surface area may increase the probability of implantation at vulnerable endometrial sites. These associations were weaker than those observed for placenta previa and intrauterine surgery and should be interpreted cautiously.

Because placenta previa is a strong and prenatally identifiable risk factor, we conducted stratified analyses by placenta previa status. Among women with neither prior CS nor placenta previa, independent risk factors were similar to those in the overall non-CS population, including operative hysteroscopy, uterine curettage, IVF, and multifetal pregnancy, aligning with prior meta-analysis[20]. In women without prior CS but with placenta previa, operative hysteroscopy and uterine curettage remained independently associated with PAS. Some investigators proposed deficient decidualization in the lower uterine segment, and PAS was triggered compounded by endometrial injury[21], which was consisted with our results. These findings suggest that placenta previa status may modify the clinical risk profile. Prenatal identification of placenta previa, combined with careful evaluation of intrauterine surgical history, could improve risk stratification and guide monitoring.

The overall model showed good discrimination (AUC 0.838), but this performance was largely influenced by the strong contribution of placenta previa. After stratification by placenta previa status, the discriminative ability was lower, particularly among women without prior CS but with placenta previa (AUC 0.647). Therefore, the model should not be viewed as a stand-alone diagnostic tool. Instead, it may help identify women who could benefit from targeted PAS ultrasound examination, referral to experienced imaging specialists, or closer antenatal review. The high NPV in the no-previa subgroup suggests potential value for ruling out PAS in low-risk settings, whereas the higher PPV in the previa subgroup may help prioritize specialist assessment; however, these findings require external validation.

These results add cohort-based evidence to previous studies of individual PAS risk factors by evaluating a defined population of women without prior CS and by reporting model discrimination, calibration, decision curve analysis, and internal validation. The value of the model lies in its transparency and use of routinely available variables. Its clinical role should be considered supportive and preliminary: it may assist triage for targeted imaging but should not be used to make definitive diagnostic or management decisions without ultrasound assessment and clinical expertise.

Stratified analysis by placenta previa status suggested that prior intrauterine surgeries remain clinically relevant beyond placenta previa or cesarean history. Nevertheless, the attenuated performance of the subgroup models indicates that clinical history alone is insufficient for refined risk prediction in some patients, especially those with placenta previa.

Future studies should prioritize multicenter external validation and should evaluate whether adding ultrasound features, serum biomarkers, or other routinely available clinical variables improves prediction. Artificial intelligence-driven approaches may be useful if supported by standardized multicenter data, but such approaches require careful validation and transparent reporting. Women with PAS without prior CS may have focal or subtle endometrial injury and less specific sonographic findings than post-cesarean cases; therefore, risk models may be most useful when integrated with specialist imaging pathways.

This study has several strengths. First, it focused on women without prior CS, a group that represents a clinically relevant but less emphasized PAS population. Second, it used a large single-center delivery cohort including 11,148 women and 236 PAS cases diagnosed according to the 2019 FIGO clinical and/or histopathological criteria. Third, it classified intrauterine surgical history by type and frequency, which may improve clinical risk assessment. Fourth, it evaluated model performance using discrimination, calibration, decision curve analysis, and stratified 5-fold cross-validation, and it reported placenta previa-stratified results to clarify model performance in clinically distinct subgroups.

This study also has important limitations. First, it was conducted at a single tertiary referral center. Peking University Third Hospital manages a relatively high proportion of complex obstetric cases referred from other hospitals, which may have increased the observed PAS prevalence and may limit generalizability to lower-risk settings. Second, although the overall cohort was large, some subgroup analyses had limited sample sizes, particularly for less common characteristics such as uterine malformations. Third, the pathological confirmation rate was relatively low, likely because women without prior CS had fewer cases of placenta percreta and a low hysterectomy rate. However, PAS was diagnosed according to FIGO 2019 clinical and/or histopathological criteria, and all hysterectomy cases were histologically confirmed. Fourth, the model was internally validated only; external validation in independent and multicenter cohorts is required before broad clinical implementation. Finally, the model was based on clinical history and placenta previa status without ultrasound features, which limits its discriminative ability in some subgroups.

## Conclusions

In women without prior CS, placenta previa, uterine curettage, operative hysteroscopy, IVF, and multifetal pregnancy were independently associated with PAS in this single-center cohort. A clinical-history-based prediction model showed good discrimination in the overall cohort but weaker discrimination after stratification by placenta previa status. The model may support preliminary risk stratification and selection of patients for targeted PAS imaging or specialist assessment, but external validation and multimodal model development are needed before routine clinical use.

## Data Availability

The datasets used and/or analysed during the current study are available from the corresponding author on reasonable request.

## Acknowledgments

The authors gratefully acknowledge the staff of Department of Obstetrics and Gynecology, Peking University Third Hospital for their support and assistance.

## Supporting information captions

**Supplemental Figure 1** Internal validation of prediction models by 5-fold cross validation.Group of patients (a)without prior CS. (b)with neither prior CS nor placenta previa.(c)with neither prior CS nor placenta previa.

**Supplemental Table 1** Baseline characteristics in placenta accreta spectrum (PAS) patients with neither prior cesarean section (CS) nor placenta previa and non-PAS controls.

**Supplemental Table 2** Multivariate analysis of risk factors for placenta accreta spectrum (PAS) patients with neither prior cesarean section (CS) nor placenta previa.

**Supplemental Table 3** Baseline characteristics in placenta accreta spectrum (PAS) patients without prior cesarean section (CS) but with placenta previa and non-PAS controls.

**Supplemental Table 4** Multivariate analysis of risk factors for placenta accreta spectrum (PAS) patients without prior cesarean section (CS) but with placenta previa.

**Supplemental Table 5** Summary of diagnostic performance metrics of three predictive models in this study.

